# Retrospective Analysis of Blood Biomarkers of Neurological Injury in Human Cases of Viral Infection and Bacterial Sepsis

**DOI:** 10.1101/2024.01.05.24300906

**Authors:** Maggie L. Bartlett, Heather Goux, Linwood Johnson, Kevin L. Schully, Melissa Gregory, Joost Brandsma, Josh G. Chenoweth, Danielle V. Clark, Luis Felipe Rivera, Carlos Lezcano-Coba, Amy Y. Vittor, Ronald Hayes, Josefrancisco Galué, Jean-Paul Carrera, Darci R. Smith

## Abstract

**Background:** Blood biomarkers of neurological injury could provide a rapid diagnosis of central nervous system (CNS) injury caused by infections. An FDA-approved assay for mild traumatic brain injury (TBI) measures glial fibrillary acidic protein (GFAP) and ubiquitin carboxy-terminal hydrolase L1 (UCH-L1), which signal astrocyte and neuronal injury, respectively. Here, we assessed the applicability of this biomarker assay for determining infection-induced brain injury.

**Methods:** We measured serum levels of GFAP and UCH-L1 retrospectively in serum samples from three study populations: 1) human cases infected with Venezuelan equine encephalitis virus (VEEV) and Madariaga virus (MADV) (n = 73), 2) human sepsis patients who were severely ill or diagnosed with encephalitis (n = 66), and 3) sepsis cases that were subsequently evaluated for cognitive impairment (n = 64).

**Results:** In the virus infection group, we found elevated GFAP for VEEV (p = 0.014) and MADV (p = 0.011) infections, which correlated with seizures (p = 0.006). In the bacterial sepsis group, GFAP was elevated in cases diagnosed with encephalitis (p = 0.0007) and correlated with headaches (p = 0.0002). In the bacterial sepsis cases with a later cognitive assessment, elevated GFAP (p = 0.0057) at study enrollment was associated with cognitive impairment six months later with a positive prognostic capacity of 79% (CI: 66–95%; p = 0.0068).

**Conclusions:** GFAP and UCH-L1 levels measured using an FDA-approved assay for TBI may indicate brain injury resulting from viral or bacterial infections and could predict the development of neurological sequelae.

## Background

Central nervous system (CNS) infections, which can lead to brain injury via invasion of brain tissue, inflammation, and/or the release of toxins, can be diagnostically challenging (1). Diagnosis requires a combination of clinical assessment, magnetic resonance imaging (MRI) studies and computed tomography (CT) scans, cerebrospinal fluid (CSF) analysis, and identification of the causative pathogen. Neurological injury biomarkers could provide a new approach to the diagnosis of infection-induced CNS injury. The first FDA-approved test for a blood biomarker for acute traumatic brain injury (TBI) detects glial fibrillary acidic protein (GFAP) and ubiquitin carboxy-terminal hydrolase L1 (UCH-L1), which signal astrocyte and neuronal injury, respectively (2). GFAP and UCH-L1 are also used as biomarkers for neurodegenerative diseases in blood or CSF, like multiple sclerosis (3, 4), neuromyelitis optica spectrum disorder (5), Alzheimer’s disease (6–8), and less often for CNS infections (9, 10).

Venezuelan equine encephalitis virus (VEEV), eastern equine encephalitis virus (EEEV), and Madariaga virus (MADV) are important mosquito-borne, single-stranded positive-sense RNA viruses (*Togaviridae*; *Alphavirus*) that circulate throughout the Americas and infect the CNS of humans and equids (11). VEEV enzootic/endemic subtype ID infection is prevalent in the eastern province of Darién, Panama, with seroprevalence up to 75% in some villages(11). EEEV was reclassified as two different species in 2010: EEEV in North America and MADV in Latin America (12). MADV was not associated with human outbreaks until 2010 when the first human MADV outbreak was reported in Darién, Panama (13). A force-of-infection model applied to two serosurveys from Darién Province in 2012 and 2017 found endemic alphavirus transmission (14). Alphavirus disease typically presents as a self-limited febrile illness but can be fatal, and persistent neurological symptoms have occurred for up to five years following MADV or VEEV exposure (11).

Sepsis, including septicemia, pneumonia, skin infections, and intra-abdominal infections, is a serious and potentially life-threatening consequence of the exaggerated release of cytokines and other inflammatory mediators in response to infection. The inflammatory response leads to cellular injury and organ dysfunction, including the CNS (15). Although bacteria can cause meningitis and brain abscesses, CNS injury may result indirectly from cytokine-mediated alterations in the blood-brain barrier (BBB), hypoxia, and microcirculatory abnormalities (13). Sepsis-associated encephalopathy affects about 30% of patients with sepsis and is characterized by an altered level of consciousness (16). Sepsis patients who recover often suffer long-term cognitive impairment resulting from BBB disruption, neuroinflammation, neurotransmitter dysfunction, and neuronal loss (17, 18).

Here, we determined the efficacy of an FDA-approved Brain Trauma Indicator (BTI) biomarker assay, designed for brain injury in mild TBI patients (2), in detecting brain injury caused by infections. The three study populations we evaluated included acute alphavirus cases, sepsis cases, and sepsis cases that were evaluated after recovery for cognitive impairment (Table 1). We measured higher levels of GFAP in all cohorts compared to uninfected controls, suggesting that GFAP is a good indicator of infection-induced brain injury and may predict neurological sequelae, such as cognitive impairment. This is the first report that supports the use of an FDA-approved blood-based biomarker assay to detect possible neurological injury in viral and bacterial infections.

**Table 1.** Study populations for neurological injury biomarker assessment.

| Group | Subgroup | Serum (N) | Country | Reference |
| --- | --- | --- | --- | --- |
| **1—Virus Infection** | Acute MADV | 8 | Panama | N/A |
|  | Acute VEEV | 22 |  |  |
|  | Both (MADV/VEEV) | 3 |  |  |
|  | Controls<sup>1</sup> | 40 |  |  |
| **2—Bacterial Infection (Sepsis)** | CNS Infection | 21 | Ghana n=10,<br>Cambodia n=11 | 20, 21 |
|  | Other Infection | 32 | Ghana n=14,<br>Cambodia n=18 |  |
|  | Controls<sup>2</sup> | 13 | Ghana | 21 |
| **3—Bacterial Infection (Sepsis with Cognitive Assessment)** | Study Enrollment (day 0) | 64 | Ghana | 21 |
|  | 28 Days Post Enrollment | 38 |  |  |
|  | Controls<sup>2</sup> | 13 |  |  |

<sup>1</sup>Group 1 controls are samples from the region that were negative for VEEV/MADV by all assays.
<sup>2</sup>Group 2 and 3 controls were healthy individuals from the region.

## Methods

### Study Populations and Sample Collection Procedures

#### Study Population 1: virus infection group

Sera and metadata were collected from consenting adults and children with parental consent during an acute alphaviral outbreak in 2018-2019 in Aruza, Darién, by the Gorgas Memorial Institute, with negative control serum samples collected from healthy individuals in the region. Sera were deidentified and shipped on dry ice to the Naval Medical Research Command (NMRC). The studies were approved by the Panamanian Ministry of Health (protocol: 2077), the Gorgas Institutional Review Board (IRB) (protocol: 335/CBI/ICGES/21), and the University of Florida IRB (protocol: 201500292).

Enzyme-linked immunosorbent assays (ELISAs) were used to measure IgM and IgG antibodies to MADV and VEEV and samples were run in duplicate. ELISA antigens were prepared from the EEEV (prepared by Dr. Robert Shope at the Yale Arbovirus Research Unit in August 1989) and VEEV complex strain 78V-3531 (from an infected mouse brain) (11). All samples were also tested for viruses using virus-specific plaque reduction neutralization tests (PRNTs). A positive sample was reported as the reciprocal of the highest dilution that reduced plaque counts by >80% (PRNT_80_). Reactivity to VEEV (strain TC-83) and MADV (strain GML-267113, isolated from a fatal human case in Panama in 2017) was measured (14).

#### Study population 2: bacterial sepsis

Sera from suspected human sepsis cases were collected by NMRC’s Austere environments Consortium for Enhanced Sepsis Outcomes (ACESO) program during prospective observational studies in Cambodia and Ghana (19, 20) under IRB-approved protocols (protocols: NMRC.2013.0019 and NMRC.2016.0004-GHA) in compliance with all applicable U.S. Federal regulations governing the protection of human subjects. All subjects, or their legally authorized representatives, provided written informed consent and authorized future use of their specimens and data. Responses were recorded on a standardized case report form and included demographics, medical history, physical exam findings, and admitting diagnosis. A diagnosis of a CNS infection was based on the judgment of two physicians. If these two physicians did not agree on a diagnosis, a third physician reviewed patient records and made the diagnosis. Other infections were confirmed by culture or polymerases chain reaction (PCR) (supplementary data 1) and included respiratory tract infection, bacteremia, abscess, cellulitis, gastrointestinal tract infection, fungemia, oral cavity infection, reproductive tract infection, or cardiovascular infection (20).

#### Study population 3: bacterial sepsis with cognitive assessment

Sera from human sepsis cases were collected by the NMRC ACESO program during prospective observational studies in Ghana as described above for study population 2. Sera were collected at enrollment (day 0; n = 64) or day 28 after enrollment (day 28; n = 38). Impairment was assessed using the Montreal Cognitive Assessment Basic (MoCA-B) (21) for individuals at enrollment (n = 64), upon discharge (n = 64), day 28 (n = 38), and months 6 (n = 32) and 12 (n = 20) after discharge, as available.

### GFAP and UCH-L1 Assays

Sera was evaluated with the Banyan BTI assay according to the manufacturer’s guidelines. Antibodies were vortexed, diluted in antibody diluent, and kit standards prepared following a serial dilution (UCH-L1: 2,560 pg/mL to 80 pg/mL; GFAP 320 pg/mL to 10 pg/mL). Serum was added to plates (UCH-L1: 25 µL; GFAP: 50 µL) in duplicate and incubated for 60 minutes at 37 °C with shaking at 500 rpm. Plates were washed, detection antibody added and allowed to incubate for 60 minutes at 37 °C while shaking at 500 rpm. Plates were washed and 150 µL of substrate added per well prior to reading optical density at 225 nm. If the concentration exceeded the upper limit of quantitation, serum was diluted and re-run. Samples that were below the lower limit of quantitation (LLOQ) were included at half the minimum LLOQ for statistical analyses. The cutoff for positivity for mild TBI in the BTI assay is 327 pg/mL UCH-L1 and 22 pg/mL GFAP.

### Statistical Analyses

Statistical analyses were performed using SPSS (Version 19.0. Armonk, NY: IBM Corp) or GraphPad v9 and included the Mann-Whitney U test following normality testing, the Jonckheere–Terpstra (JT) test, and area under the curve plots. A chi-square test was used to determine statistical significance for age and sex differences. The relevant tests are indicated in figure legends.

## Results

### Study Population 1: Virus Infection

For population 1, serum samples were collected from 33 cases of acute alphavirus infection in eastern Panama. Subjects with confirmed infections tested positive for VEEV, MADV, or both by IgM ELISA and PRNT (Supplemental Table 1), whereas subjects with likely infection tested positive for either IgM ELISA or PRNT. Symptoms consistent with alphavirus illness included fever, chills, headaches, and, less commonly, myalgia, arthralgia, nausea, vomiting, and seizures (Figure 1A, Table 2). Patients infected with MADV had a higher incidence of seizures (37.5%) compared to those infected with VEEV ID (5.0%), and the only fatality occurred in an individual with MADV. Serum levels of GFAP and UCH-L1 were determined with the BTI assay for uninfected controls and the alphavirus-positive individuals. Mean UCH-L1 levels were significantly lower in cases of VEEV but not in cases of MADV, whereas the mean GFAP levels were significantly higher for subjects with either alphavirus (Figure 1B). However, UCH-L1 was elevated in the fatal case and in two out of the four cases with seizures. The highest levels of GFAP were observed in three of the four cases with seizures and the fatal case. When grouped by time since onset of fever (early 0–7 days, mid 8–14 days, or late >15 days), UCH-LI decreased (p = 0.00031) during the mid-timeframe and was statistically unchanged either early or late (Figure 1C). However, GFAP was elevated at the early (p = 0.013) and mid (p = 0.0059) timeframes but not late (p = 0.092) (Figure 1C), which may be due to the small sample size at this time point. For symptoms, seizures were significantly associated with higher levels of GFAP (p = 0.006) (Table 2). Interestingly, GFAP levels were higher in those without inflammatory response symptoms, including fever, chills, and myalgia, and this increase was statistically significant for chills (p = 0.023). There were no significant associations between levels of UCH-L1 and symptoms. A Wilcoxon rank-sum analysis indicated that the relationship between GFAP and acute alphavirus infection, i.e., infection with both VEEV and MADV, acute MADV infection, or acute VEEV infection was strengthened, while the relationship between UCH-L1 and any infection remained unchanged (Supplemental Table 2). These results suggest that GFAP may be a promising biomarker of acute alphavirus disease and may indicate disease severity.

**Figure 1.**
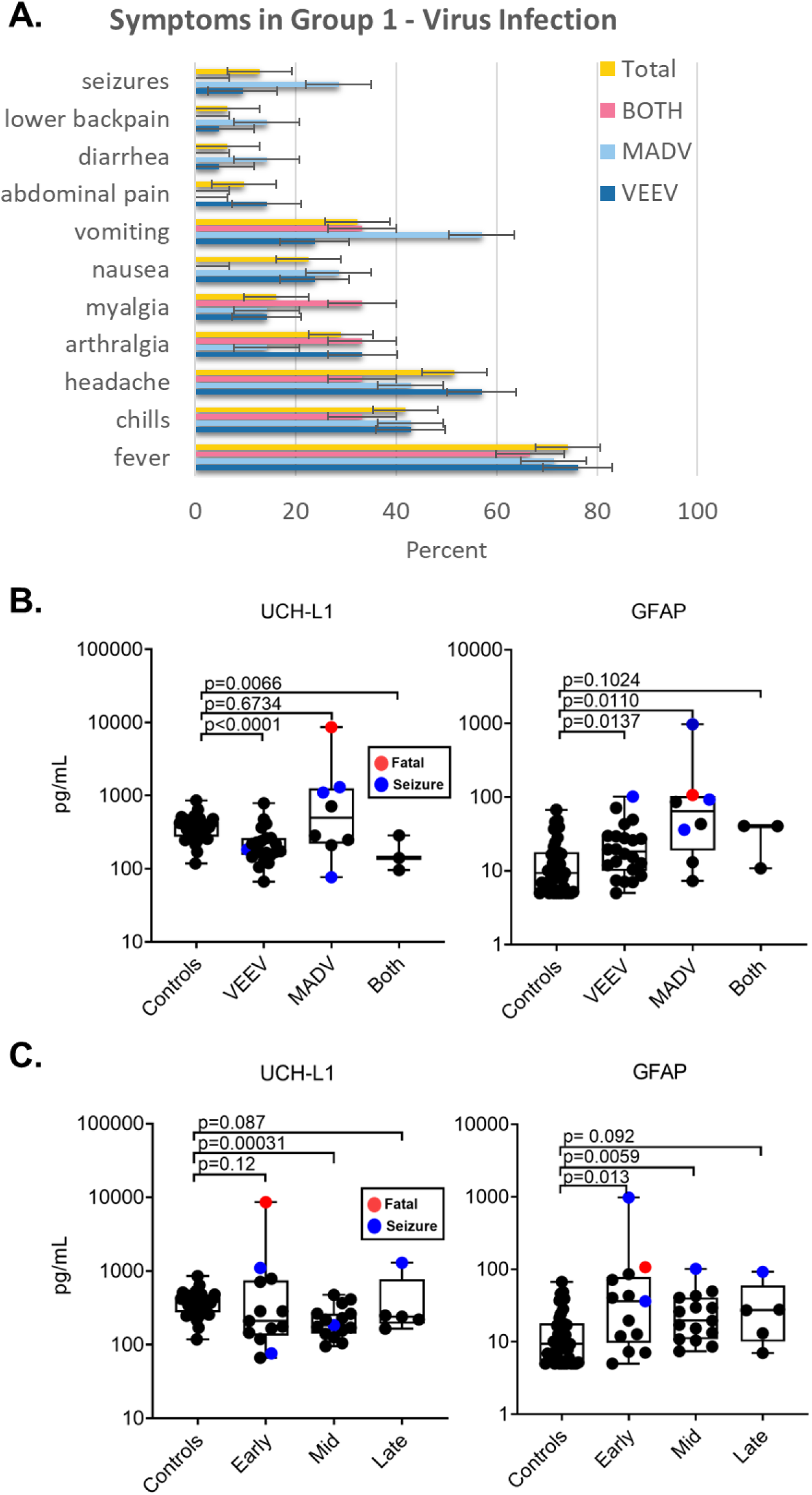
Neurotropic alphavirus exposure cases in Panama from the group 1 virus infection study population. A) (n = 31) were grouped based on antibody reactivity to MADV (n = 7), VEEV (n = 21), and both (n = 3). B) UCH-L1 and GFAP levels (pg/mL) in patient serum (n = 33), where each point represents a single patient: MADV (n = 8), VEEV (n = 22), both (n = 3) and negative (n = 40). Blue points represent patients with seizures (n = 4), and red indicates mortality (n = 1). Comparison of analyte levels in patients grouped by positive or negative for any virus and days of clinical evolution: early (0–7), mid (8–14), late (>15) for C) UCH-L1 and GFAP (pg/mL). Blue points represent patients with seizures (n = 4), and red indicates mortality (n = 1). Statistical significance is based on Mann-Whitney U test vs. negative controls for both viruses.

**Table 2.** Associations between symptoms and biomarkers for group 1 virus infection study population. Clinical symptoms reported in 30 cases for which sex was reported. Statistical significance was determined by a chi-square test.

|  |  | N | GFAP pg/ml (95% CI) | p-value | UCH-L1 pg/ml (95% CI) | p-value |
| --- | --- | --- | --- | --- | --- | --- |
| Sex | Male | 22 | 70.4 (-20.4, 161.2) | 0.482 | 281.4 (143.2, 419.6) | 0.159 |
|  | Female | 8 | 38.0 (13.8, 62.2) |  | 369.3 (159.0, 579.7) |  |
| Age (mean) | 0-10 | 8 | 157.6 (-121.1, 436.2) | 0.881 | 494.8 (109.0, 880.6) | 0.306 |
|  | 11-20 | 8 | 25.4 (2.8, 48.0) |  | 260.7 (103.5, 418.0) |  |
|  | 21-30 | 6 | 18.8 (6.7, 30.9) |  | 188.3 (134.6, 242.0) |  |
|  | 31-40 | 5 | 28.4 (-22.6, 79.3) |  | 160.7 (76.5, 244.8) |  |
|  | >41 | 3 | 44.7 (-19.8, 109.3) |  | 389.5 (-483.7, 1262.8) |  |
| Fever | Yes | 23 | 33.4 (20.9, 46.0) | 0.391 | 295.3 (173.0, 417.6) | 0.902 |
|  | No | 7 | 154.8 (-182.0, 491.5) |  | 336.3 (7.2, 665.4) |  |
| Chills | Yes | 13 | 20.4 (7.0, 33.8) | 0.023 | 222.5 (128.1, 316.9) | 0.277 |
|  | No | 17 | 93.4 (-25.0, 211.7) |  | 367.9 (182.5, 553.2) |  |
| Headache | Yes | 16 | 22.9 (11.7, 34.1) | 0.135 | 243.5 (161.8, 325.2) | 0.868 |
|  | No | 14 | 106.2 (-40.1, 252.5) |  | 375.0 (145.9, 604.1) |  |
| Arthralgia | Yes | 9 | 19.8 (4.2, 35.5) | 0.08 | 232.1 (69.3, 394.9) | 0.205 |
|  | No | 21 | 79.7 (-15.1, 174.5) |  | 336.0 (188.5, 483.6) |  |
| Myalgia | Yes | 5 | 16.4 (6.7, 26.1) | 0.303 | 195.1 (125.4, 264.7) | 0.636 |
|  | No | 25 | 70.8 (-8.24, 149.9) |  | 326.8 (195.1, 458.5) |  |
| Nausea | Yes | 7 | 38.0 (8.9, 67.1) | 0.508 | 328.7 (128.7, 528.7) | 0.447 |
|  | No | 23 | 69.0 (-17.6, 155.6) |  | 297.6 (159.5, 435.7) |  |
| Vomiting | Yes | 10 | 40.2 (16.8, 63.6) | 0.482 | 382.5 (118.3, 646.7) | 0.218 |
|  | No | 20 | 72.5 (-28.0, 173.0) |  | 266.0 (147.9, 384.2) |  |
| Abdominal pain | Yes | 3 | 32.7 (-52.2, 117.6) | 0.863 | 357.1 (-569.6, 1283.7) | 0.809 |
|  | No | 27 | 65.0 (-8.12, 138.1) |  | 299.1 (182.9, 415.2) |  |
| Diarrhea | Yes | 2 | 25.1 (-204.9, 255.1) | 0.678 | 224.6 (-534.6, 983.7) | 0.868 |
|  | No | 28 | 64.4 (-6.01, 134.7) |  | 310.6 (192.4, 428.8) |  |
| Lower back pain | Yes | 1 | 17.1 | 0.862 | 229.8 | 0.773 |
|  | No | 29 | 623.3 (-4.6, 131.1) |  | 307.4 (193.3, 421.6) |  |
| Seizures | Yes | 4 | 302.6 (-417.4, 1022.7) | 0.006 | 665.5 (-329.0, 1659.9) | 0.428 |
|  | No | 26 | 24.7 (16.3, 33.1) |  | 249.4 (179.1, 319.6) |  |

### Study Population 2: Bacterial Sepsis

We analyzed patient samples from sepsis studies completed in Cambodia (20) and Ghana (19) to measure changes in UCH-L1 or GFAP as a result of bacterial infection. This study population was comprised of 53 severely ill, hospitalized patients who presented with at least two systemic inflammatory response syndrome criteria and a suspected infection. These included a sepsis cohort with symptoms consistent with CNS disease (n = 21) and individuals without symptoms of CNS disease but positive for infection (n = 32). A control group (n = 13) comprised healthy individuals from the same region. A diagnosis of a CNS infection was determined after a review by two physicians of the patient’s medical history, which included evaluation for nerve palsies, paralysis, reflex response abnormalities, and assessment by the Glasgow coma scale. The severely ill patients presented with fever, headache, abdominal pain, nausea, and diarrhea (Figure 2A). This cohort reported more myalgia than the alphavirus cohort; however, there was no difference in myalgia between CNS and other infections. Serum levels of GFAP and UCH-L1 were determined with the BTI assay for control samples, the severely ill individuals with no signs of a CNS infection, and those diagnosed with a CNS infection. We found no significant differences in UCH-L1 levels; however, GFAP levels were significantly higher in individuals with CNS infections and elevated, although not significantly, in patients with non-CNS infections (Figure 2B). Also, patients with fatal vs. non-fatal cases had similar levels of UCH-L1 and GFAP. Among symptoms, headache was positively associated with GFAP (p = 0.0002) but negatively associated with UCH-L1 (p < 0.0001) (Table 3). These results are similar to those for our viral infection study population (group 1) and indicate that elevated levels of GFAP may be a potential biomarker to indicate CNS infection.

**Figure 2.**
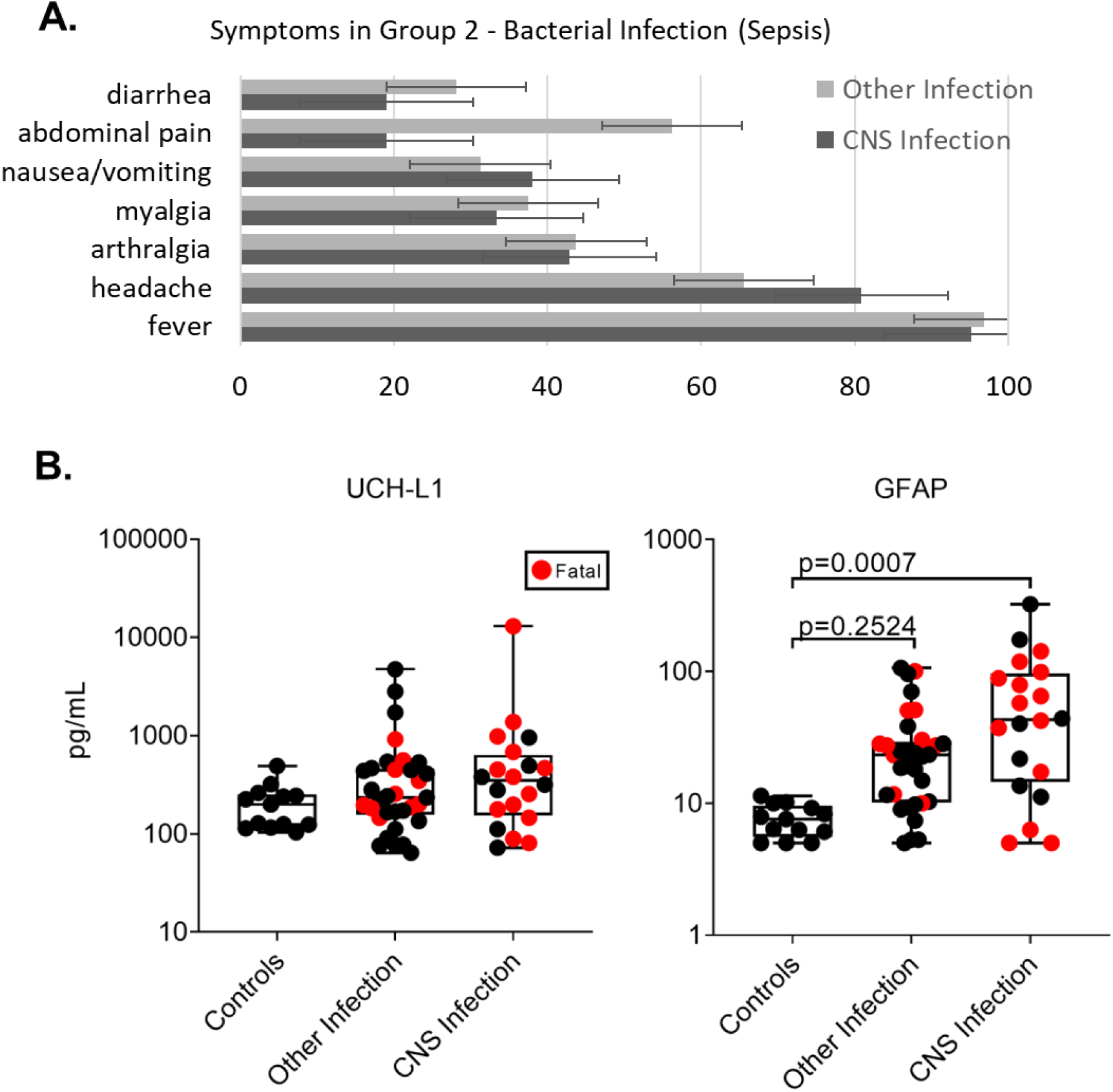
Severe, hospitalized acute febrile illness cases in Ghana and Cambodia with or without CNS involvement in the group 2 bacterial sepsis cohort. A) Patient-reported symptoms were grouped based on the clinical designation of CNS infection determined by two physicians. B) UCH-L1 and GFAP levels (pg/mL) grouped by CNS, other, or controls. Statistical significance is based on comparison biomarker values at corresponding time points using Mann-Whitney U test vs. healthy controls.

**Table 3.**
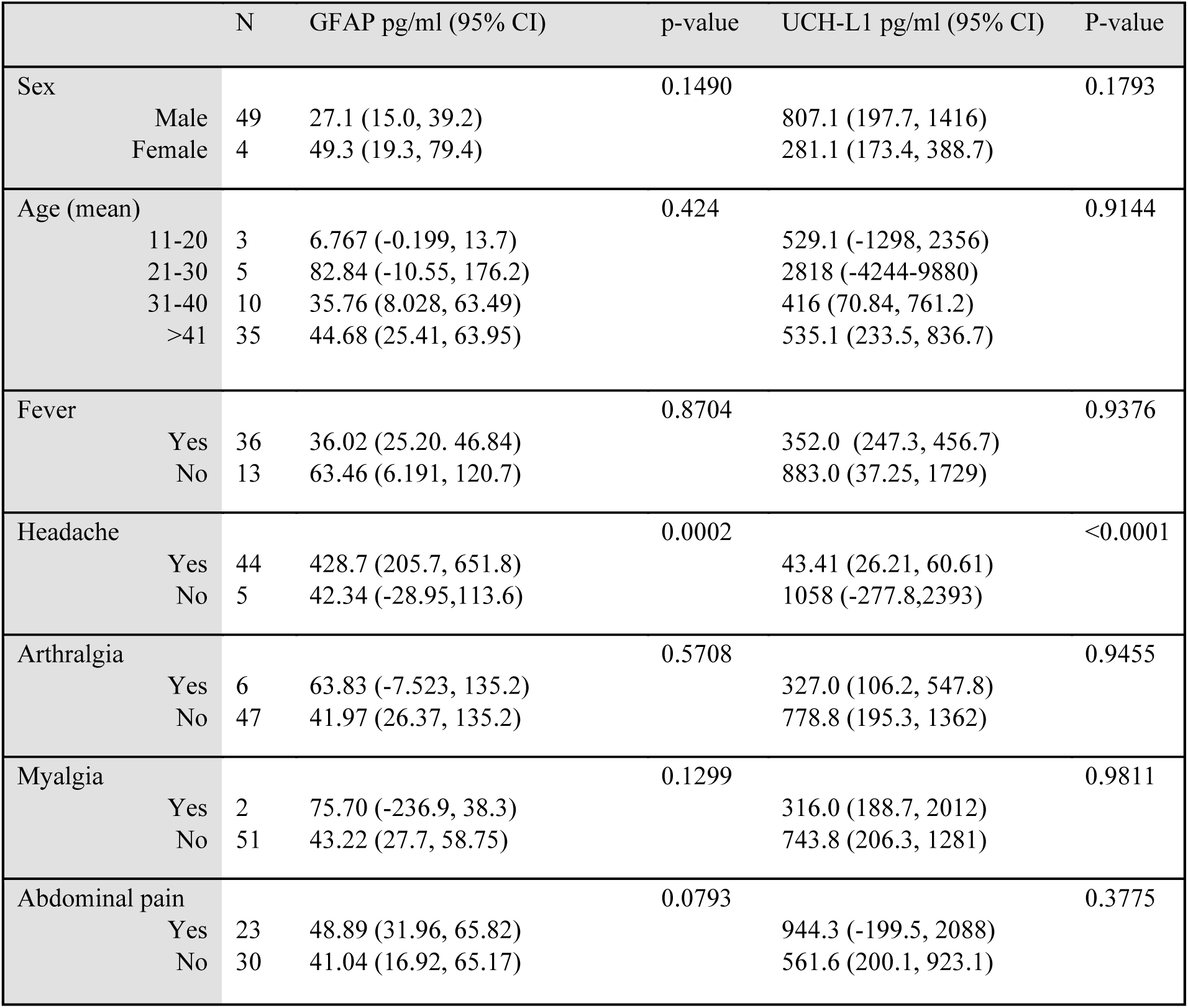
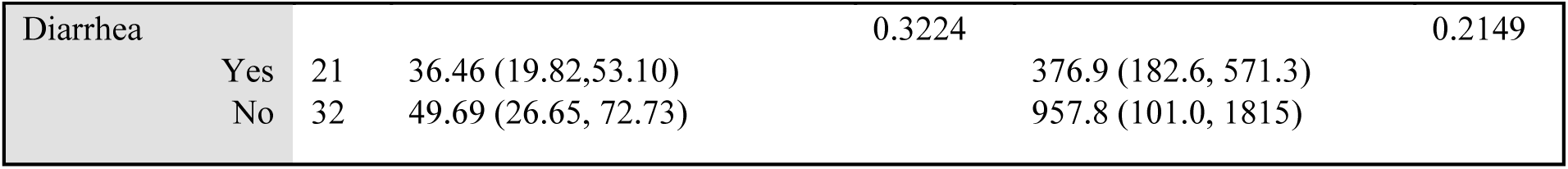
Associations between symptoms and biomarkers in group 2 bacterial infection (sepsis). Statistical significance was determined by a chi-square test.

### Study Population 3: Bacterial Sepsis with Cognitive Assessment

Finally, we determined whether the levels of these biomarkers measured at study enrollment would predict cognitive impairment after sepsis based on MoCA-B scores, which were measured during study enrollment and on subsequent visits up to a year later. MoCA-B scoring differentiates among cognitive syndromes by assessing memory, executive function, attention, language, visuospatial parameters, and orientation (21). A normal score is ≥ 26, whereas impairment falls into mild (18–25), moderate (10–17), and severe (< 10) categories. Previous sepsis studies completed in Ghana (19) provided serum samples collected prospectively from hospitalized cases of sepsis at study enrollment (day 0) and day 28 after enrollment. MoCA-B scores were determined for individuals at study enrollment, upon discharge, on day 28, and 6 and 12 months after discharge, when feasible. Levels of UCH-L1 and GFAP were measured in individuals at study enrollment (day 0) and day 28 after enrollment. Mean UCH-L1 levels were unchanged, whereas the mean GFAP levels were significantly higher at both time points (Figure 3A). We then evaluated the levels of GFAP and UCH-L1 at study enrollment (day 0) stratified by the average MoCA score after discharge and up to 12 months later. Levels of UCH-L1 trended higher in cases with severe cognitive impairment and persisted up to 12 months after symptom onset; however, they did not reach statistical significance (Figure 3B). Levels of GFAP in serum at study enrollment increased with increasing levels of cognitive impairment vs. controls up to twelve months after study enrollment (Figure 3B). The percentage of individuals who developed cognitive impairment after discharge from the hospital was compared to the results of the BTI assay at study enrollment (Table 4). Individuals with a higher level of cognitive impairment after discharge were more likely to have had a positive BTI assay at study enrollment. Thus, the Banyan BTI assay may predict the development of reduced cognitive function after infection. Supporting these associations, trends for UCH-L1 (p = 0.00363) and GFAP (p = 1.849 × 10^−8^) were significant by the JT test for ranked categories. Thus, serum GFAP levels at study enrollment predicted cognitive impairment as measured by MoCA-B scores at six months. The six-month time point was chosen for comparison because the sample size was split equally between patients with normal (≥26) or impaired (<26) MoCA-B scores. Not only was cognitive impairment associated with higher levels of GFAP at enrollment (p = 0.0057), but this metric should identify 79% (CI: 66–95%; p=0.0068) of cases correctly based on time-dependent receiver operating characteristic (ROC) curves (Figure 3C and Figure 3D).

**Figure 3.**
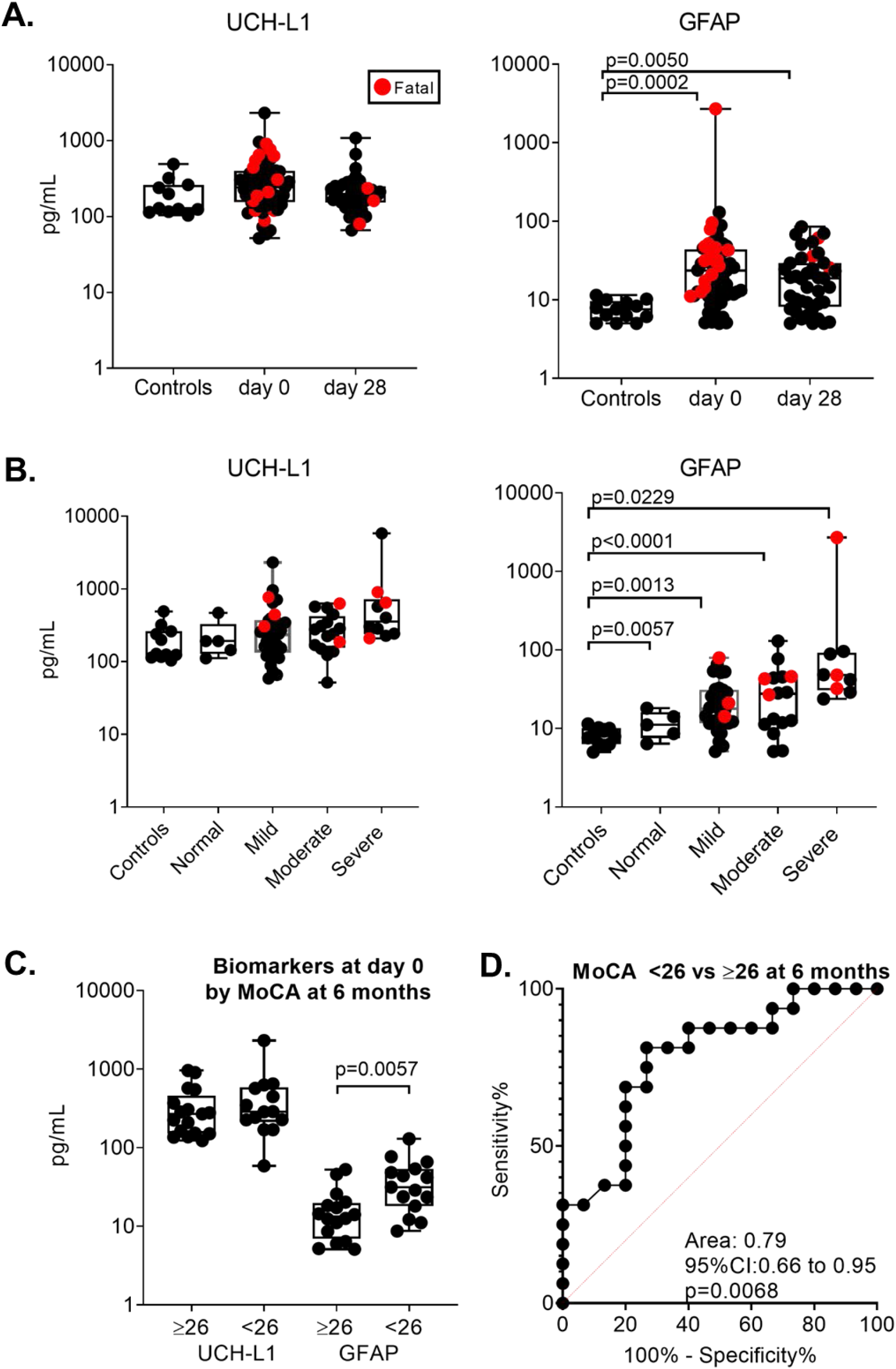
Longitudinal study of group 3 severe bacterial sepsis with cognitive assessment. A) UCH-L1 and GFAP levels (pg/mL) in patient sera from Ghana and Cambodia grouped by study enrollment (day 0) or day 28 post enrollment compared to controls. B) UCH-L1 and GFAP (pg/mL) levels in sera from patients grouped by the average MoCA severity score at discharge and up to six months later. Statistical significance is based on values at corresponding time points using Mann-Whitney U test vs. controls. No MoCA surveys were obtained from controls. C) Comparison of UCH-L1 and GFAP between patients with normal (≥26) or impaired (<26) MoCA severity scores at six months post enrollment. D) AUC analysis of the predictive value of GFAP at study enrollment to impaired cognitive function six months later.

**Table 4.** Cognitive impairment on follow up in group 3 bacterial infection (sepsis with cognitive assessment).

| MoCA category | Cognitive impairment after discharge percentage (n) | BTI positive at enrollment percentage (n) |
| --- | --- | --- |
| Normal ($\geq 26$) | 8.2% (5) | 20% (1) |
| Mild (25-18) | 49.2% (30) | 46.7 % (14) |
| Moderate (17-10) | 26.2% (16) | 56.3% (9) |
| Severe ($<10$) | 16.4% (9) | 100% (9) |

Therefore, GFAP levels may serve as an effective biomarker for predicting the development of cognitive impairment in individuals with severe infectious diseases.

## Discussion

We evaluated two brain biomarkers using an FDA-approved BTI assay developed originally for acute TBI, in which elevation of either or both biomarkers are considered indicative of brain injury (2). GFAP is an intermediate filament protein expressed almost exclusively in astrocytes, where it is induced by neural injury and released upon disintegration of the astrocyte cytoskeleton (22). UCH-L1 is expressed only in neurons where it is involved in the ubiquitination of proteins intended for degradation by the proteasomal pathway and plays an important role in the removal of oxidized or misfolded proteins in both normal and pathological conditions (8). Analyzing both GFAP and UCH-L1 may reflect distinct causes of brain injury. In severe TBI, focal mass lesions cause an increase in GFAP, whereas diffuse brain injuries cause an increase in UCH-L1 (23, 24). Here, we found a significant increase in GFAP, but not UCH-L1, in the alphavirus and sepsis cohorts, suggesting that GFAP is a shared biomarker for brain injury and infectious diseases of the CNS.

Previous work on biomarkers, including GFAP, analyzed only the CSF, not serum from viral encephalitis cases, and none used an FDA-approved assay (25–30). Early identification of sepsis-associated encephalopathy, one of the most serious clinical manifestations of sepsis (31), would provide a more accurate diagnosis and prognosis, which now relies on nonspecific clinical manifestations and imaging features, often followed by lumbar punctures and electroencephalograms. Several biomarkers have been proposed for the rapid detection of sepsis-related brain injury (32), including serum GFAP and UCH-L1, which correlate with the occurrence, severity, and prognosis of sepsis-associated encephalopathy (33, 34). We found higher levels of GFAP in acute CNS-involved sepsis, and these higher levels of serum GFAP at the time of hospitalization correlated with cognitive decline up to twelve months after symptom onset in our group 3 cohort. We also observed elevated UCH-L1, similar to the previous study (33), but it was not statistically significant, which may be due to the sample size or time of sampling. Therefore, GFAP and possibly UCH-L1 may serve as biomarkers for sepsis-related brain injury and should be further evaluated.

Lesions in the brain of fatal human VEEV cases appear to result from injury due to virus replication and clearance of infected cells by the immune system (35). Although neurons are the primary targets of encephalitic alphaviruses and viral cytopathology is important in CNS dysfunction, fatal encephalitis is likely mediated by the immune response to infection rather than direct virus replication (36). For individuals infected by the encephalitic alphaviruses, mean levels of GFAP and the incidence of seizures were significantly higher than controls, whereas the mean UCH-L1 levels were reduced significantly in VEEV but not MADV. The latter results were unexpected because neurons are the major target cells of the encephalitic alphaviruses, but these results may be due to the time of sampling. In cell culture and animal models, encephalitic alphaviruses also infect astrocytes (37–39), which are immuno-activated in infected animals (36). Our GFAP results likely indicate that astrocytes were injured by virus infection or the immune response.

Our results suggests that GFAP levels may serve as an effective biomarker for predicting the development of cognitive impairment in individuals with severe infectious diseases. Neurological sequelae after recovery from alphavirus infection occurs frequently and includes seizures, paralysis, intellectual disability, behavioral changes, photophobia, coma, or visual disturbances (11, 40). GFAP may predict the development of neurologic sequelae after alphavirus infection, similar to our results with the bacterial sepsis cohort, and is a focus of our future studies.

In conclusion, CNS infections have high rates of morbidity and mortality and blood-based diagnostic assays for brain injury could aid clinicians when deciding whether imaging is needed and to initiate treatment. Our data support the use of the BTI assay to identify brain injury caused by CNS infection that could lead to neurological sequelae. Further research is needed to identify the relevant timeframes, examine additional cohorts, and conduct clinical trials to determine the impact of this assay on patient outcomes. Our study lays the foundation for these efforts.

## Data Availability

All data produced in the present work are contained in the manuscript.

## Authors’ contributions

ML Bartlett, DR Smith, and A Vittor had full access to the study data and take full responsibility for the integrity and accuracy of this analysis. Concept and design: ML Bartlett, DR Smith, A Vittor, R Hayes; Data acquisition, analysis, and interpretation: ML Bartlett, DR Smith, A Vittor, R Hayes, JP Carrera, LF Rivera, C Lezcano-Coba, J Galue, KL Schully, M Gregory, J Brandsma, JG Chenoweth, DV Clark; Laboratory analysis: DR Smith, J Linwood, H Goux, JP Carrera, C Lezcano-Coba and J Galue; Drafting of the manuscript: ML Bartlett, DR Smith; KL Schully, JG Chenoweth, R Hayes, JP Carrera; Statistical analysis: ML Bartlett, A Vittor, J Brandsma.

## Acknowledgments

This study was supported by the Defense Threat Reduction Agency under project number CB10937 to DRS and grant number FID-2021-96 SENACYT to JPC. The views expressed in this work reflect the results of research conducted by the authors and do not necessarily reflect the official policy or position of the Department of Defense, the Navy, or the United States Government. Several of the authors are U.S. Government employees. This work was prepared as part of their official duties. Title 17, U.S.C., § 105 provides that copyright protection under this title is not available for any work of the U.S. Government. Title 17 U.S.C., § 101 defines a U.S. Government work as work prepared by a military Service member or employee of the U.S. Government as part of that person’s official duties.

