## Supplemental Tables for "Retrospective Analysis of Blood Biomarkers of Neurological Injury in Human Cases of Viral Infection and Bacterial Sepsis"

Supplemental Table 1. Human cases of acute alphavirus infection for group 1 virus infection study population.

| Virus | | MADV Previous Exposure[3] | MADV Naïve[4] | VEEV Previous Exposure[3] | VEEV Naïve[4] | Total |
|---|---|---|---|---|---|---|
| **Both** | MADV and VEEV Confirmed[1] | N/A | N/A | N/A | N/A | 0 |
| | VEEV Confirmed[1] and MADV Likely[2] | N/A | N/A | N/A | N/A | 2 |
| | MADV and VEEV Likely[2] | N/A | N/A | N/A | N/A | 1 |
| **MADV** | MADV Confirmed[1] | N/A | N/A | 1 | 1 | 2 |
| | MADV Likely[2] | N/A | N/A | 3 | 3 | 6 |
| **VEEV** | VEEV Confirmed[1] | 15 | 2 | N/A | N/A | 17 |
| | VEEV Likely[2] | 2 | 3 | N/A | N/A | 5 |

[1]Confirmed infection determined by IgM and PRNT positivity.

[2]Likely infection determined by either IgM or PRNT positivity, but not both.

[3]Previous exposure determined by IgG positivity.

[4]Naive indicates no antibodies detected.

Supplemental Table 2. Associations between group 1 virus infection subgroups and biomarkers.

| Protein | Infection | Adj variance | z | P-value |
|---|---|---|---|---|
| GFAP | Acute alphavirus | 8119.16 | -3.474 | 0.0005 |
| | Acute MADV | 3875.05 | -2.940 | 0.0033 |
| | Acute VEEV | 6511.54 | -1.834 | 0.0513 |
| UCH-L1 | Acute alphavirus | 8139.75 | 3.486 | 0.0005 |
| | Acute MADV | 3884.88 | -0.610 | 0.5421 |
| | Acute VEEV | 7535.43 | 4.810 | <0.0001 |

Wilcoxon rank-sum tests of GFAP/UCHL1 and alphaviral status
