## Supplemental Data 1 for "Retrospective Analysis of Blood Biomarkers of Neurological Injury in Human Cases of Viral Infection and Bacterial Sepsis"

**Supplementary Data 1: Group 2 and 3**

| ID | Original Diagnosis | Additional Diagnosis | Country | Group(s) |
|---|---|---|---|---|
| 01C01 008 | CNS Infection | None | Cambodia | 2 |
| 01C01 015 | CNS Infection | None | Cambodia | 2 |
| 01C01 038 | CNS Infection | None | Cambodia | 2 |
| 01C01 062 | CNS Infection | None | Cambodia | 2 |
| 01C01 085 | CNS Infection | None | Cambodia | 2 |
| 01C01 099 | CNS Infection | None | Cambodia | 2 |
| 01C01 107 | CNS Infection | None | Cambodia | 2 |
| 01C01 114 | CNS Infection | None | Cambodia | 2 |
| 01C01 131 | CNS Infection | None | Cambodia | 2 |
| 01C01 175 | CNS Infection | None | Cambodia | 2 |
| 01C01 195 | CNS Infection | None | Cambodia | 2 |
| 01G01 0015 | CNS Infection | None | Ghana | 2&3 |
| 01G01 0074 | CNS Infection | None | Ghana | 2 |
| 01G01 0079 | CNS Infection | None | Ghana | 2 |
| 01G01 0085 | CNS Infection | None | Ghana | 2 |
| 01G01 0108 | CNS Infection | None | Ghana | 2 |
| 01G01 0123 | CNS Infection | None | Ghana | 2 |
| 01G01 0143 | CNS Infection | None | Ghana | 2 |
| 01G01 0152 | CNS Infection | None | Ghana | 2 |
| 01G01 0153 | CNS Infection | None | Ghana | 2 |
| 01G01 0182 | CNS Infection | None | Ghana | 2 |
| 01C01 187 | Fungemia, GU or Reproductive Infection | Escherichia coli | Cambodia | 2 |
| 01C01 030 | GI Tract Infections | Escherichia coli, Proteus | Cambodia | 2 |
| 01C01 052 | GI Tract Infections | Escherichia coli, Kleb pneumo | Cambodia | 2 |
| 01C01 057 | GI Tract Infections | Orientia tsutsugamushi | Cambodia | 2 |
| 01G01 0136 | HEENT or Oral Cavity Infection | ENTEROBACTER AEROGENES | Ghana | 2 |
| 01C01 033 | Respiratory Tract Infection | Staphlococcus aureus | Cambodia | 2 |
| 01C01 045 | Respiratory Tract Infection | Orientia tsutsugamushi | Cambodia | 2 |
| 01C01 102 | Respiratory Tract Infection | Klebsiella pneumoniae | Cambodia | 2 |
| 01C01 123 | Respiratory Tract Infection | Enterobacter spp., S. aureus | Cambodia | 2 |
| 01C01 166 | Respiratory Tract Infection | Orientia tsutsugamushi | Cambodia | 2 |
| 01C01 189 | Respiratory Tract Infection | Haemophilus influenzae, Klebsiella pne | Cambodia | 2 |
| 01G01 0028 | Respiratory Tract Infection | None | Ghana | 2&3 |
| 01C01 111 | Unknown Source/Infection | Haemophilus influenzae | Cambodia | 2 |
| 01C01 168 | Unknown Source/Infection | Orientia tsutsugamushi | Cambodia | 2 |
| 01G01 0012 | unknown source/infection | None | Ghana | 2&3 |
| 01G01 0017 | unknown source/infection | None | Ghana | 2&3 |
| 01C01 152 | Bacteremia | Klebsiella pneumoniae | Cambodia | 2 |
| 01G01 0026 | Bacteremia | SALMONELLA SPP. ISOLATED | Ghana | 2 |
| 01G01 0086 | Bacteremia | METHICILLIN-RESISTANT STAPHYL | Ghana | 2&3 |
| 01G01 0050 | Bacteremia | None | Ghana | 2&3 |
| 01C01 054 | Bacteremia AND Unknown Source/Infection | Orientia tsutsugamushi | Cambodia | 2 |
| 01C01 089 | Bacteremia, Cellulitis/Abscess/Skin/bone/ Joint Infection | Staphlococcus aureus | Cambodia | 2 |
| 01G01 0048 | Bacteremia, Cellulitis/Abscess/Skin/bone/ Joint Infection | None | Ghana | 2&3 |
| 01G01 0175 | Bacteremia, Cellulitis/Abscess/Skin/bone/ Joint Infection | STREPTOCOCCUS PYOGENES ISOL | Ghana | 2&3 |
| 01G01 0039 | Bacteremia, GU or Reproductive Infection | E-COLI | Ghana | 2&3 |
| 01G01 0070 | Bacteremia, GU or Reproductive Infection | STAPH AUREUS | Ghana | 2 |
| 01C01 183 | Bacteremia, HEENT or Oral Cavity Infection, Respiratory T | Klebsiella pneumoniae, Pseudomonas | Cambodia | 2 |
| 01G01 0034 | Bacteremia, Respiratory Tract Infection | STREPTOCOCCUS PNEUMONIAE | Ghana | 2 |
| 01G01 0008 | bacterimia | None | Ghana | 2 |
| 01C01 072 | Cardiovascular Infection | Streptococcus pneumoniae | Cambodia | 2 |
| 01G01 0139 | Cardiovascular Infection, Cellulitis/Abscess/Skin/bone/ Join | STREPTOCOCCUS PNEUMONIAE | Ghana | 2 |
| 01G01 0083 | Cellulitis/Abscess/Skin/bone/ Joint Infection | None | Cambodia | 2 |
| 01G01000200hEP0# | Non-Infectious | Unknown | Ghana | 3 |
| 01G01000500hEP0# | GI Tract Infections | Unknown | Ghana | 3 |
| 01G01000700hEP0# | GI Tract Infections | Unknown | Ghana | 3 |
| 01G01000800hEP0# | Bacteremia | Unknown | Ghana | 3 |
| 01G01001000hEP0# | Non-Infectious | Unknown | Ghana | 3 |
| 01G01001100hEP0# | Cellulitis/Abscess/Skin/bone/ Joint Infection | Unknown | Ghana | 3 |
| 01G01001800hEP0# | Parasitemia | Unknown | Ghana | 3 |
| 01G01001900hEP0# | Cellulitis/Abscess/Skin/bone/ Joint Infection | Unknown | Ghana | 3 |
| 01G01002000hEP0# | Cellulitis/Abscess/Skin/bone/ Joint Infection | Unknown | Ghana | 3 |
| 01G01002500hEP0# | Parasitemia | Unknown | Ghana | 3 |
| 01G01002700hEP0# | Cellulitis/Abscess/Skin/bone/ Joint Infection | Unknown | Ghana | 3 |
| 01G01002900hEP0# | HEENT or Oral Cavity Infection | Unknown | Ghana | 3 |
| 01G01003300hEP0# | GI Tract Infections, Respiratory Tract Infection | Unknown | Ghana | 3 |
| 01G01003800hEP0# | Respiratory Tract Infection | Unknown | Ghana | 3 |
| 01G01004400hEP0# | GI Tract Infections | Unknown | Ghana | 3 |
| 01G01004500hEP0# | Respiratory Tract Infection | Unknown | Ghana | 3 |
| 01G01005100hEP0# | Cellulitis/Abscess/Skin/bone/ Joint Infection | Unknown | Ghana | 3 |

| | | | | |
|---|---|---|---|---|
| 01G01005300hEP0# | Unknown Source/Infection | Unknown | Ghana | 3 |
| 01G01005400hEP0# | Non-Infectious | Unknown | Ghana | 3 |
| 01G01005800hEP0# | Non-Infectious | Unknown | Ghana | 3 |
| 01G01006000hEP0# | Cellulitis/Abscess/Skin/bone/ Joint Infection | Unknown | Ghana | 3 |
| 01G01006100hEP0# | Respiratory Tract Infection | Unknown | Ghana | 3 |
| 01G01006400hEP0# | Cellulitis/Abscess/Skin/bone/ Joint Infection | Unknown | Ghana | 3 |
| 01G01006500hEP0# | Respiratory Tract Infection | Unknown | Ghana | 3 |
| 01G01006800hEP0# | GI Tract Infections, GU or Reproductive Infection | Unknown | Ghana | 3 |
| 01G01007100hEP0# | Non-Infectious | Unknown | Ghana | 3 |
| 01G01007300hEP0# | GI Tract Infections | Unknown | Ghana | 3 |
| 01G01007500hEP0# | GU or Reproductive Infection | Unknown | Ghana | 3 |
| 01G01007800hEP0# | Cellulitis/Abscess/Skin/bone/ Joint Infection | Unknown | Ghana | 3 |
| 01G01008300hEP0# | GU or Reproductive Infection | Unknown | Ghana | 3 |
| 01G01008900hEP0# | Non-Infectious | Unknown | Ghana | 3 |
| 01G01009300hEP0# | Cellulitis/Abscess/Skin/bone/ Joint Infection | Unknown | Ghana | 3 |
| 01G01009500hEP0# | GU or Reproductive Infection | Unknown | Ghana | 3 |
| 01G01009600hEP0# | GU or Reproductive Infection | Unknown | Ghana | 3 |
| 01G01010500hEP0# | Bacteremia, GU or Reproductive Infection | Unknown | Ghana | 3 |
| 01G01011100hEP0# | Parasitemia | Unknown | Ghana | 3 |
| 01G01011500hEP0# | Cellulitis/Abscess/Skin/bone/ Joint Infection | Unknown | Ghana | 3 |
| 01G01012500hEP0# | Respiratory Tract Infection | Unknown | Ghana | 3 |
| 01G01012800hEP0# | Respiratory Tract Infection | Unknown | Ghana | 3 |
| 01G01012900hEP0# | Unknown Source/Infection | Unknown | Ghana | 3 |
| 01G01013200hEP0# | Cellulitis/Abscess/Skin/bone/ Joint Infection | Unknown | Ghana | 3 |
| 01G01014400hEP0# | Parasitemia | Unknown | Ghana | 3 |
| 01G01014700hEP0# | Bacteremia, GU or Reproductive Infection | Unknown | Ghana | 3 |
| 01G01014900hEP0# | Parasitemia | Unknown | Ghana | 3 |
| 01G01015400hEP0# | Parasitemia | Unknown | Ghana | 3 |
| 01G01015700hEP0# | Respiratory Tract Infection | Unknown | Ghana | 3 |
| 01G01015800hEP0# | Cellulitis/Abscess/Skin/bone/ Joint Infection | Unknown | Ghana | 3 |
| 01G01015900hEP0# | Bacteremia | Unknown | Ghana | 3 |
| 01G01016300hEP0# | Cellulitis/Abscess/Skin/bone/ Joint Infection | Unknown | Ghana | 3 |
| 01G01016900hEP0# | Cellulitis/Abscess/Skin/bone/ Joint Infection | Unknown | Ghana | 3 |
| 01G01017200hEP0# | Parasitemia | Unknown | Ghana | 3 |
| 01G01017400hEP0# | GU or Reproductive Infection | Unknown | Ghana | 3 |
| 01G01017800hEP0# | Cellulitis/Abscess/Skin/bone/ Joint Infection | Unknown | Ghana | 3 |
| 01G01018300hEP0# | Cellulitis/Abscess/Skin/bone/ Joint Infection , Parasitemia | Unknown | Ghana | 3 |
| 01G01019200hEP0# | Respiratory Tract Infection | Unknown | Ghana | 3 |
| 01G01 9029 | Healthy Control | Not applicable | Ghana | 2&3 |
| 01G01 9030 | Healthy Control | Not applicable | Ghana | 2&3 |
| 01G01 9031 | Healthy Control | Not applicable | Ghana | 2&3 |
| 01G01 9032 | Healthy Control | Not applicable | Ghana | 2&3 |
| 01G01 9033 | Healthy Control | Not applicable | Ghana | 2&3 |
| 01G01 9034 | Healthy Control | Not applicable | Ghana | 2&3 |
| 01G01 9035 | Healthy Control | Not applicable | Ghana | 2&3 |
| 01G01 9036 | Healthy Control | Not applicable | Ghana | 2&3 |
| 01G01 9037 | Healthy Control | Not applicable | Ghana | 2&3 |
| 01G01 9038 | Healthy Control | Not applicable | Ghana | 2&3 |
| 01G01 9040 | Healthy Control | Not applicable | Ghana | 2&3 |
| 01G01 9045 | Healthy Control | Not applicable | Ghana | 2&3 |
| 01G01 9046 | Healthy Control | Not applicable | Ghana | 2&3 |
